# Auto-Detection and Segmentation of Involved Lymph Nodes in HPV-Associated Oropharyngeal Cancer Using a Convolutional Deep Learning Neural Network

**DOI:** 10.1101/2022.01.19.22269566

**Authors:** Nicolette Taku, Kareem A. Wahid, Lisanne V. van Dijk, Jaakko Sahlsten, Joel Jaskari, Kimmo Kaski, C. David Fuller, Mohamed A. Naser

## Abstract

**Purpose:** Segmentation of involved lymph nodes on head and neck computed tomography (HN-CT) scans is necessary for the radiotherapy treatment planning of human papilloma virus (HPV) associated oropharynx cancers (OPC). We aimed to train a deep learning convolutional neural network (DL-CNN) to identify and segment involved lymph nodes on contrast-enhanced HN-CT scans.

**Methods:** 90 patients who underwent levels II-IV neck dissection for newly diagnosed, clinically node-positive, HPV-OPC were identified. Ground-truth segmentation of all radiographically and pathologically involved nodes was manually performed on pre-surgical HN-CT scans, which were randomly divided into training/validation dataset (n=70) and testing dataset (n=20). A 5-fold cross validation was used to train 5 separate DL-CNN sub-models based on a residual U-net architecture. Validation and testing segmentation masks were compared to ground-truth segmentation masks using overlap-based, volume-based, and distance-based metrics. A lymph auto-detection model was developed by thresholding segmentation model outputs, and 20 node-negative HN-CT scans were added to the test set to further evaluate auto-detection capabilities. Model discrimination of lymph node “positive” and “negative” HN-CT scans was evaluated using the area under the receiver operating characteristic curve (AUC).

**Results:** In the DL-CNN validation phase, all sub-models yielded segmentation masks with median DSC ≥ 0.90 and median volume similarity score of ≥ 0.95. In the testing phase, the DL-CNN produced consensus segmentation masks with median Dice of 0.92 (IQR, 0.89-0.95), median volume similarity of 0.97 (IQR, 0.94-0.99), and median Hausdorff distance of 4.52 mm (IQR, 1.22-8.38). The detection model achieved an AUC of 0.98.

**Conclusion:** The results from this single-institution study demonstrate the successful automation of lymph node segmentation for patients with HPV-OPC using a DL-CNN. Future studies, including external validation using a larger dataset, are necessary to clarify the role of the DL-CNN in the routine radiation oncology treatment planning workflow.

## Introduction

Approximately 66,000 cases of head and neck cancer will be diagnosed in the United States in 2021, including 30% of cases pertaining to human papilloma virus (HPV)-associated oropharynx cancers (OPC) ^1,2^. Accurate assessment of the extent of lymph node involvement and lymph node characteristics on staging studies is necessary for appropriate treatment disposition. Some patients with early-stage HPV-associated OPC, including limited lymph node involvement and no radiographic evidence of extranodal extension, can be managed with transoral robotic resection of the primary site of disease and ipsilateral neck dissection. However, the majority of patients diagnosed with locoregionally advanced disease will receive radiotherapy treatment with definitive intent, thereby necessitating imaging-based segmentation of the primary tumor and involved lymph nodes to ensure adequate radiotherapy dose delivery to all sites of disease ^3^.

The acquisition of head and neck computed tomography (HN-CT) scans for HPV-associated OPC is an integral component of primary tumor and nodal staging as well as radiotherapy treatment planning. Several studies have demonstrated unique imaging characteristics for HPV-associated OPC ^4,5^. In a blinded, matched-pair analysis of HN-CT scans for patients with HPV-positive and HPV-negative OPC, Cantrell et al. found that HPV-positive OPC scans were less likely to demonstrate muscle invasion of the primary tumor but more likely to demonstrate cystic morphology of involved lymph nodes ^6^. Similarly, Chan et al. observed that HPV-positive OPC was more likely to demonstrate multiple lymph node involvement and cystic nodal appearance ^7^. These unique radiographic features correspond to histopathology findings observed on the surgical specimens of HPV-associated OPC tumors ^8^.

Deep learning is a subset of machine learning that uses deep neural networks to learn and classify data ^9^. Within the context of OPC, deep learning algorithms have been used to predict HPV status based on pre-treatment imaging ^10,11^. Although clinical assessment of involved lymph nodes is necessary for therapy disposition and radiotherapy treatment planning, no deep learning algorithms have focused on the identification and segmentation of involved lymph nodes for HPV-associated OPC. The purpose of this study was to develop a deep learning convolutional neural network (DL-CNN) capable of identifying and segmenting radiographically and pathologically involved lymph nodes for HPV-associated OPC on contrast-enhanced HN-CT scans. Furthermore, we aimed to use the DL-CNN to discriminate between node-negative and node-positive HN-CT scans.

## Methods

After obtaining Institutional Review Board approval, 90 patients who underwent selective, levels II-IV neck dissection for newly diagnosed, clinically node-positive, OPC at our institution were identified from the Steifel Oropharynx Database—a prospective database of clinical and patient-reported outcomes for patients treated at The University of Texas MD Anderson Cancer Center. In addition, 20 randomly selected patients who underwent selective, levels II-IV neck dissection and were found to have clinically and pathologically node-negative OPC were included in the dataset. The inclusion criterion were at least 18 years of age at the time of diagnosis and pathology findings consistent with HPV-associated OPC, while the exclusion criteria were a history of radiotherapy treatment to the head and neck region or a history of prior neck dissection.

### Data Preparation and Preprocessing

Pre-surgical, contrast-enhanced, HN-CT scans were identified for all patients. Expert, “ground-truth segmentation” of all radiographically involved lymph nodes was manually performed on node-positive HN-CT scans using RayStation Research (RaySearch Laboratories, Stockholm, Sweden) ^12^. Histopathology findings from selective neck dissection were correlated with neuroradiology annotations to ensure that 1) all segmented lymph nodes corresponded to pathologically involved lymph nodes and 2) no radiographically occult lymph nodes were present on surgical pathology. The ground-truth segmentations for each patient were then combined into a solitary “ground-truth mask”.

Pre-processing was performed on HN-CT scans to mitigate the variabilities in image size and resolution. The images and structure files were converted from Digital Imaging and Communications in Medicine (DICOM) format to Neuroimaging Informatics Technology Initiative (NIfTI) format using the Advanced Medical Imaging Registration Engine (ADMIRE, Elekta AB, Stockholm, Sweden). The images were cropped to a specific sub-volume, with the auto-segmented cephalad border of the mandible, the manually-segmented cephalad border of the sternum, and the auto-segmented external patient contour serving as the superior, inferior, and circumferential boundaries, respectively (**Figure 1**). Image intensities were then truncated to the range of [−100, 300] Hounsfield units and rescaled to the range of [-1, 1] to increase soft tissue contrast ^13^. The images and their respective ground-truth masks were resampled to 1.0 mm isotropic resolution using a trilinear interpolator in ADMIRE.

**Figure 1:**
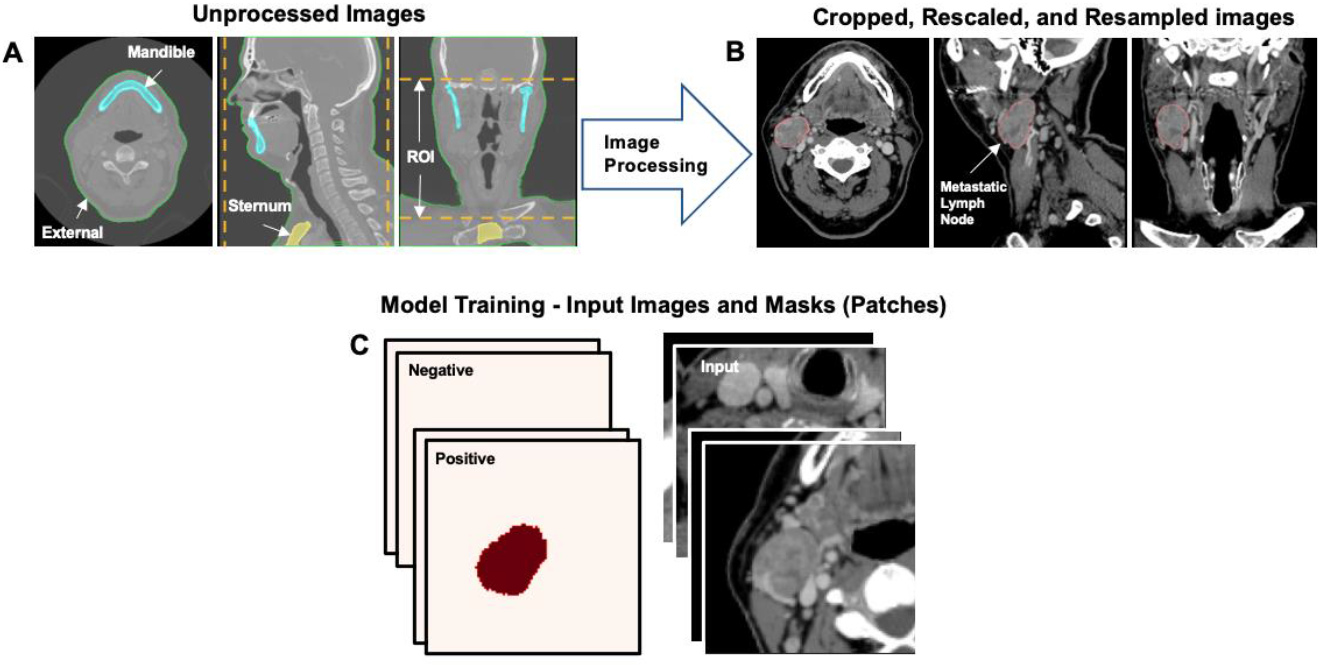
Schematic representation pre-processing workflow. Head and neck computed tomography scans were cropped using the mandible, sternum, and external contours as boundaries (A & B). Scans were divided into 4 patches of 96 × 96 × 96 voxels in dimension (C).

### Model Development

A DL-CNN was developed based on a 3-dimensional (3D) residual U-Net architecture included in the Medical Open Network for Artificial Intelligence (MONAI) software package ^14^. This architecture has been utilized successfully in previous OPC tumor auto-segmentation studies ^15^. The network consisted of 4 convolution blocks in the encoding and decoding branches with a bottleneck convolution block separating these two branches (**Figure 2**). In the encoding branch, all convolutional layers used a kernel size of 3, with each block consisting of a two-strided convolution layer; the residual connections contained a two-strided and one-strided convolution layer. In the decoding branch, all convolutional layers used a kernel size of 3, with each block consisting of a two-strided transpose convolution layer, a one-strided convolution layer, and a residual connection. In the bottleneck, all convolutional layers used a kernel size of 1 and the residual connection consisted of a two-strided convolution layer. Throughout the architecture, we utilized batch normalization and Parametric Rectified Linear Unit (PReLU) activation functions.

**Figure 2:**
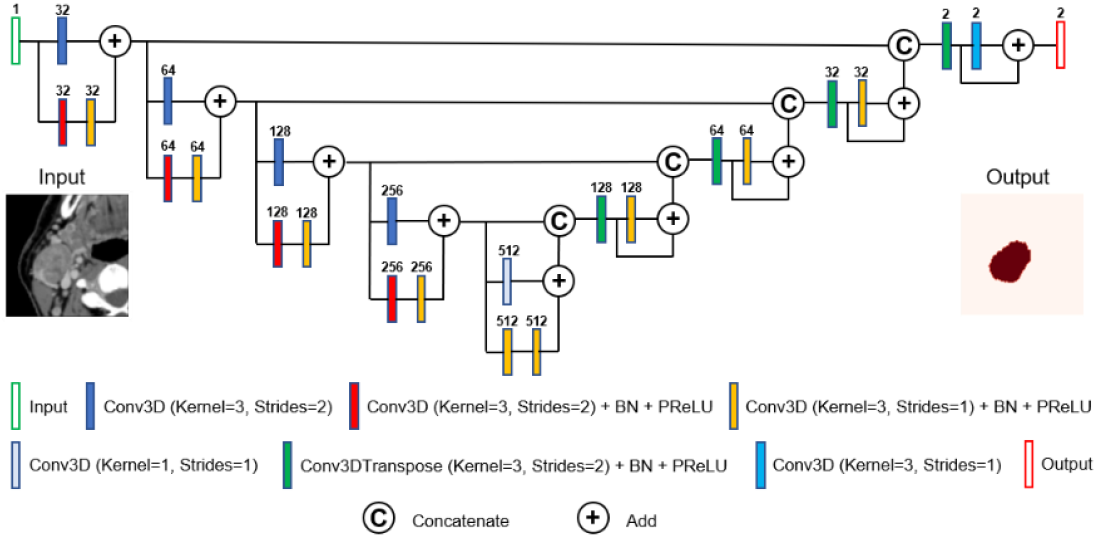
Schematic representation of the U-Net architecture implemented for the deep learning convolutional neural network with annotations pertaining to the number of channels, batch normalization (BN) layers, and Parametric Rectified Linear Unit (PReLU) layers.

### Model Training & Validation

The 90, node-positive HN-CT scans and their respective ground-truth masks served as the data by which the DL-CNN was developed. The data were randomly divided into 2 datasets—a training/validation dataset (n=70) and a testing dataset (n=20). Each HN-CT scan was split into four, random regions (i.e., patches) of 96 × 96 × 96 voxels in dimension. The input tensor consisted of a batch size of 2, a single channel input, and 4 patches per image, yielding a summative input of (8, 1, 96, 96, 96). Each patch was evaluated for the presence of an involved lymph node with the center as foreground (i.e., involved lymph node present) or background (i.e., involved lymph node absent) with a 50% probability for either condition. Several data augmentation processes were implemented to minimize overfitting. Random spatial cropping was performed to patch the images and ground-truth masks. Random horizontal flips with 50% probability, and random affine transformations with an axial rotation range of 12 degrees, and scale range of 10% were also performed.

We implemented a 5-fold cross-validation approach to train 5 separate sub-models for the DL-CNN. For each of the 5 sub-models, 80% of the HN-CT scans in the training/validation dataset and their respective ground-truth masks acted as model inputs for training purposes. The remaining 20% of HN-CT scans served for internal validation. One “validation segmentation mask” was generated per HN-CT scan, for a total of 70 validation segmentation masks. Validation segmentation masks were compared to ground-truth masks using overlap-based (Dice similarity coefficient [DSC]) and volume-based (volume similarity) metrics. The DL-CNN was trained for 700 epochs, with a learning rate of 2×10^−4^ for the first 550 epochs and 1×10^−4^ for the remaining 150 epochs.

### Model Testing

The performance of the DL-CNN to detect and segment involved lymph nodes was evaluated using an independent test dataset of 20 positive HN-CT scans. Additionally, 20 randomly selected HN-CT scans pertaining to patients with no involved lymph nodes were included in the testing dataset to evaluate the ability of the model to discriminate between “positive” (i.e., involved lymph node present) and “negative” (i.e., involved lymph node absent) HN-CT scans. In total, 5 “testing segmentation masks” were generated per HN-CT scan (1 testing segmentation mask per sub-model). For the 20 node-positive scans, the 5 testing segmentation masks for each HN-CT scan were combined to create a “consensus segmentation mask” using the Simultaneous Truth and Performance Level Estimation (STAPLE) algorithm (**Figure 3**) ^16^. The testing segmentation masks and consensus segmentation masks were compared to their respective ground-truth masks using overlap-based (DSC), volume-based (volume similarity), spatial distance-based (Hausdorff distance [HD]), and probabilistic-based (Cohen Kappa Coefficient [CKC]) metrics ^17^.

**Figure 3:**
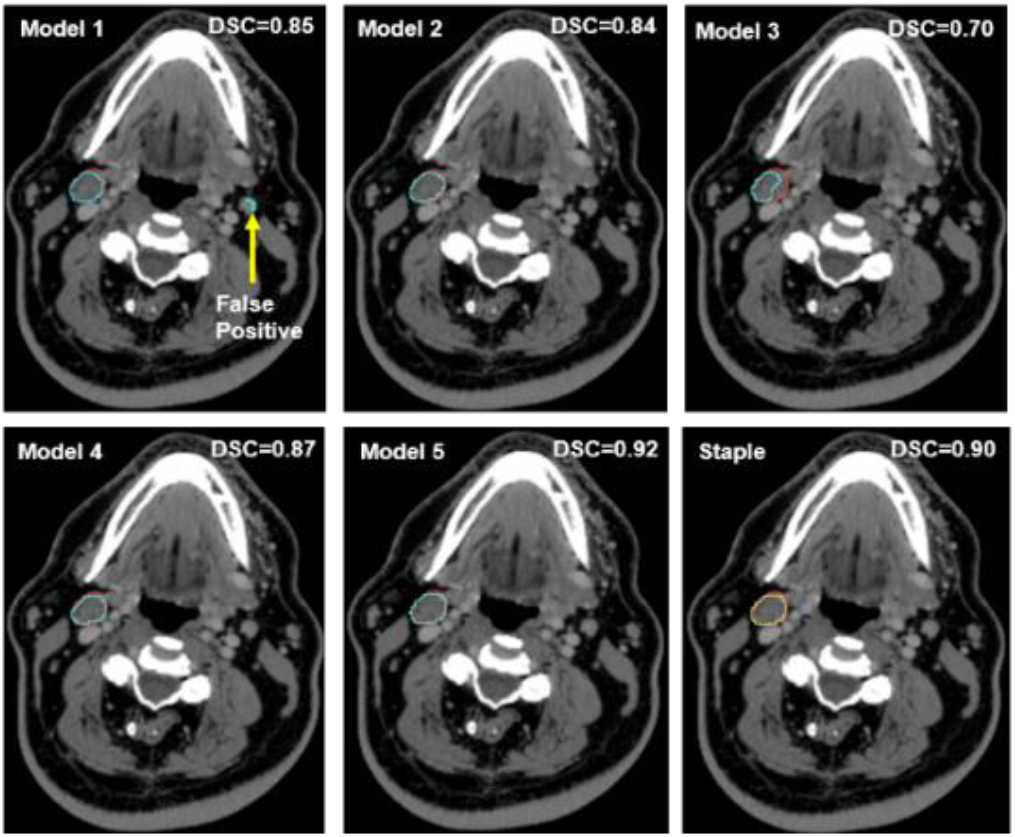
Five sub-model segmentation masks and one consensus segmentation mask were generated for each head and neck computed tomography scan. The red contour corresponds to the ground-truth masks, the blue contours correspond to the predicted sub-model segmentation masks, and the yellow contour corresponds to the consensus segmentation mask generated by combing the 5 sub-model segmentation masks using the Simultaneous Truth and Performance Level Estimation (STAPLE) algorithm.

For the model discrimination, each voxel in the 5 testing segmentation masks generated from each sub-model for the 40 HN-CT scans in the testing dataset was scored as either “1” to indicate that a lymph node contour was generated or “0” to indicate that no lymph node contour was generated. The scores for each voxel were averaged for the 5 sub-models to yield an “average score” ranging from 0 (i.e., no testing segmentation mask generated by any of the 5 sub-models) to 1 (i.e., testing segmentation masks were generated by all 5 sub-models). A HN-CT scan was considered “positive” if any voxel average score was equal to 1, and “negative” if any voxel average score was ≤ 0.8. This score threshold was chosen empirically from test results to maximize the accuracy, sensitivity, and positive predictive value of the DL-CNN. The model discrimination was evaluated by determining the area under the receiver operating characteristic curve (AUC). Three image resampling resolutions—high (1.0 mm), medium (1.5 mm), and low (2.0 mm)—were used to evaluate the impact of image resolution on the discriminatory ability of the DL-CNN.

## Results

### Patient and Tumor Characteristics

Patient and tumor characteristics are presented in **Table 1**. The median age at diagnosis was 60 years and there was a male sex predominance (n=101, 92%). The majority of the patients had no history of cigarette smoking (n=72, 66%) and cT1 disease (n=63, 57%). Among cN1 patients, there was a median of 1 involved lymph node (range, 1-4) in the training/validation dataset and 1 involved lymph node (range, 1-3) in the testing dataset.

**Table 1:** Patient and tumor clinical characteristics for all patients (N=110), patients in the training/validation dataset (n=70), and patients in the testing dataset (n=40). Abbreviations: IQR, interquartile range; y, years

| Characteristic | All<br>n (%) | Training/Validation<br>n (%) | Testing<br>n (%) |
| --- | --- | --- | --- |
| Median age [IQR], y | 60 [53-65] | 60 [54-65] | 59 [53-67] |
| Sex |  |  |  |
| Male | 101 (91.8) | 66 (94.3) | 35 (87.5) |
| Female | 9 (8.2) | 4 (5.7) | 5 (12.5) |
| Smoking Status |  |  |  |
| Never | 72 (65.5) | 44 (62.9) | 28 (70.0) |
| Former | 34 (30.9) | 24 (34.3) | 10 (25.0) |
| Current | 4 (3.6) | 2 (2.9) | 2 (5.0) |
| Oropharynx subsite |  |  |  |
| Base of tongue | 51 (46.4) | 38 (54.3) | 13 (32.5) |
| Tonsil | 59 (53.6) | 32 (45.7) | 27 (67.5) |
| Clinical tumor classification |  |  |  |
| cT1 | 63 (57.3) | 44 (62.9) | 19 (47.5) |
| cT2 | 47 (42.7) | 26 (37.1) | 21 (52.5) |
| Clinical lymph node classification |  |  |  |
| cN0 | 20 (18.2) | 0 (0.0) | 20 (50.0) |
| cN1 | 90 (81.8) | 70 (100.0) | 20 (50.0) |
| Involved lymph nodes |  |  |  |
| 0 | 20 (18.2) | 0 (0.0) | 20 (50.0) |
| 1 | 68 (61.8) | 53 (75.7) | 16 (40.0) |
| 2 | 18 (16.4) | 16 (22.9) | 2 (5.0) |
| 3 | 3 (2.7) | 0 (0) | 2 (5.0) |
| 4 | 1 (0.9) | 1 (1.4) | 0 (0.0) |

### DL-CNN Validation Performance

Segmentation mask metrics for model validation are presented in **Table 2**. When compared to ground-truth masks, sub-model #4 achieved the highest median DSC, with a score of 0.92 (interquartile range [IQR], 0.90-0.94) for the validation segmentation masks. All the 5 sub-models generated validation segmentation masks with a median DSC of at least 0.90. Similarly, all the 5 sub-models generated validation segmentation masks with a median volume similarity score of at least 0.95, with sub-model #1 achieving the highest median volume similarity score and narrowest volume similarity IQR.

**Table 2:**
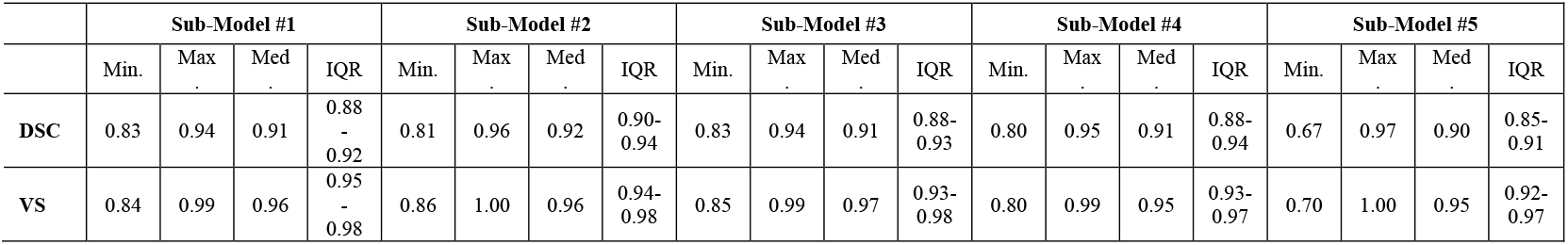
Minimum, maximum, median, interquartile range values for the overlap-based (Dice similarity coefficient) and volume-based (volume similarity) metrics for the sub-model validation segmentation masks when compared to the ground-truth masks. Abbreviations: DSC, Dice similarity coefficient; IQR, interquartile range; Max., maximum; Min., minimum; VS, volume similarity

|  | Sub-Model #1 |  |  |  | Sub-Model #2 |  |  |  | Sub-Model #3 |  |  |  | Sub-Model #4 |  |  |  | Sub-Model #5 |  |  |  |
| --- | --- | --- | --- | --- | --- | --- | --- | --- | --- | --- | --- | --- | --- | --- | --- | --- | --- | --- | --- | --- |
|  | Min. | Max. | Med. | IQR | Min. | Max. | Med. | IQR | Min. | Max. | Med. | IQR | Min. | Max. | Med. | IQR | Min. | Max. | Med. | IQR |
| **DSC** | 0.83 | 0.94 | 0.91 | 0.88-0.92 | 0.81 | 0.96 | 0.92 | 0.90-0.94 | 0.83 | 0.94 | 0.91 | 0.88-0.93 | 0.80 | 0.95 | 0.91 | 0.88-0.94 | 0.67 | 0.97 | 0.90 | 0.85-0.91 |
| **VS** | 0.84 | 0.99 | 0.96 | 0.95-0.98 | 0.86 | 1.00 | 0.96 | 0.94-0.98 | 0.85 | 0.99 | 0.97 | 0.93-0.98 | 0.80 | 0.99 | 0.95 | 0.93-0.97 | 0.70 | 1.00 | 0.95 | 0.92-0.97 |

### DL-CNN Testing Performance

Segmentation mask metrics for model testing are presented in **Table 3**. When compared to ground-truth masks, the median DSC for testing segmentation masks was greater than 0.90 for all sub-models. The median DSC for consensus segmentation masks was 0.92 (IQR, 0.89-0.95). Comparisons between the testing segmentation masks and ground-truth masks for a subset of cases based on DSC are depicted in **Figure 4**. A maximum volume similarity score of 1.0 was achieved by all sub-models for testing segmentation masks, with sub-model #4 achieving the highest minimum volume similarity score and median volume similarity score of 0.97. The median volume similarity score for consensus segmentation masks was 0.97 (IQR, 0.94-0.99). All sub-models achieved a median HD less than 6 mm, with a median HD for consensus segmentation masks of 4.52 mm (IQR, 1.22-8.38). The median CKC for testing segmentation masks was nearly identical across the sub-models, and the median CKC for consensus segmentation masks was 0.92 (IQR, 0.89-0.95).

**Table 3:** Minimum, maximum, median, interquartile range values for the overlap-based (Dice similarity coefficient), volume-based (volume similarity), spatial distance-based (Hausdorff distance), and probabilistic-based (Cohen Kappa Coefficient) metrics for the sub-model testing segmentation masks and consensus segmentation masks when compared ground-truth masks. Abbreviations: CKC, Cohen Kappa Coefficient; DSC, Dice similarity coefficient; HD, Hausdorff distance (in mm); IQR, interquartile range; Max., maximum; Min., minimum; STAPLE, Simultaneous Truth and Performance Level Estimation; VS, volume similarity

|  | Sub-Model #1 |  |  |  | Sub-Model #2 |  |  |  | Sub-Model #3 |  |  |  | Sub-Model #4 |  |  |  | Sub-Model #5 |  |  |  | Consensus (STAPLE) |  |  |  |
| --- | --- | --- | --- | --- | --- | --- | --- | --- | --- | --- | --- | --- | --- | --- | --- | --- | --- | --- | --- | --- | --- | --- | --- | --- |
|  | Min. | Max. | Med. | IQR | Min. | Max. | Med. | IQR | Min. | Max. | Med. | IQR | Min. | Max. | Med. | IQR | Min. | Max. | Med. | IQR | Min. | Max. | Med. | IQR |
| **DSC** | 0.55 | 0.95 | 0.92 | 0.89-0.94 | 0.66 | 0.95 | 0.92 | 0.88-0.94 | 0.58 | 0.95 | 0.92 | 0.87-0.94 | 0.62 | 0.96 | 0.91 | 0.87-0.94 | 0.69 | 0.96 | 0.92 | 0.88-0.94 | 0.61 | 0.96 | 0.92 | 0.89-0.95 |
| **VS** | 0.64 | 1.00 | 0.97 | 0.95-0.98 | 0.70 | 1.00 | 0.96 | 0.92-0.99 | 0.59 | 1.00 | 0.97 | 0.93-0.99 | 0.73 | 1.00 | 0.97 | 0.91-0.99 | 0.72 | 1.00 | 0.97 | 0.92-0.99 | 0.68 | 1.00 | 0.97 | 0.94-0.99 |
| **HD** | 1.11 | 92.0 | 4.92 | 1.11-16.0 | 1.22 | 90.0 | 5.78 | 1.22-17.4 | 1.65 | 86.9 | 5.08 | 1.64-18.5 | 1.22 | 90.7 | 4.15 | 1.22-9.04 | 1.22 | 91.4 | 5.56 | 1.22-11.7 | 1.22 | 90.9 | 4.52 | 1.22-8.38 |
| **CKC** | 0.55 | 0.95 | 0.92 | 0.89-0.94 | 0.66 | 0.95 | 0.92 | 0.88-0.94 | 0.58 | 0.95 | 0.92 | 0.87-0.95 | 0.62 | 0.96 | 0.91 | 0.87-0.94 | 0.69 | 0.96 | 0.92 | 0.88-0.94 | 0.61 | 0.96 | 0.92 | 0.89-0.95 |

**Figure 4:**
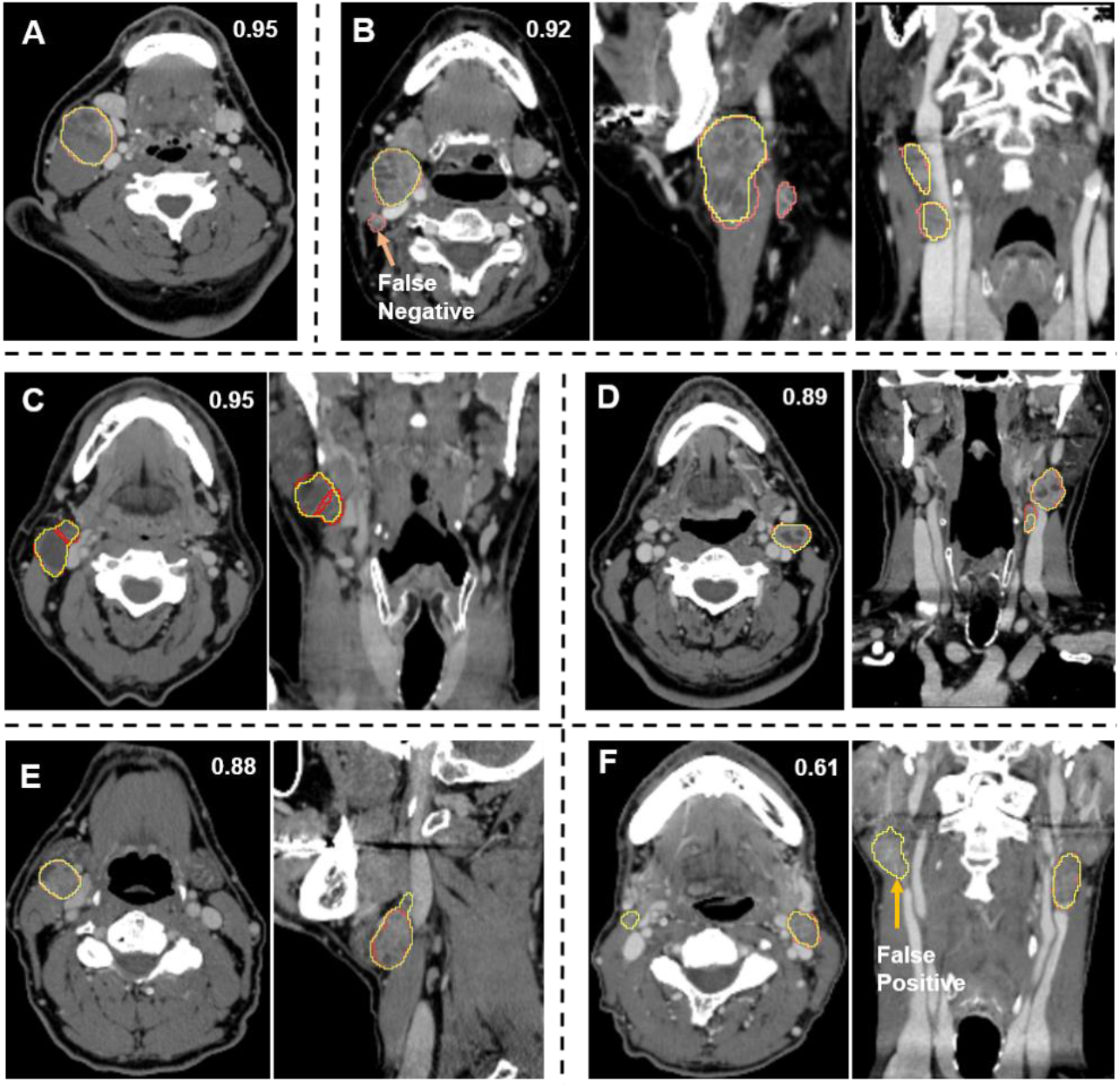
Comparison of consensus segmentations (yellow) to ground-truth segmentations (red) for a subset of test set patients with greater or equal Dice similarity coefficients (A, B, C; 1 involved lymph node, 3 involved lymph nodes, and 2 involved lymph nodes, respectively), slightly lower Dice similarity coefficients (D, E; 2 involved lymph nodes and 1 involved lymph node, respectively), and much lower Dice similarity coefficient (F; 1 involved lymph node) than the median value of 0.92.

### DL-CNN Discrimination Performance

Confusion matrices and receiver operating characteristic curves for the three imaging resolutions are presented in **Figure 5**. The medium resampled resolution model achieved the most optimal identification for the positive HN-CT scans (AUC = 0.98), with 20 of 20 HN-CT scans with involved lymph nodes correctly identified as positive and 19 of 20 of the remaining HN-CT scans correctly identified as negative. In contrast, the low resampled resolution model had the worst classification of HN-CT scans (AUC = 0.81), with 2 of 20 HN-CT scans with involved lymph nodes incorrectly identified as negative and 6 of 20 of HN-CT scans with no involved lymph nodes incorrectly identified as positive. Illustrative examples of the detection process and individual test case predictions using the best-performing model (medium resolution) are shown in **Figure S1**.

**Figure 5:**
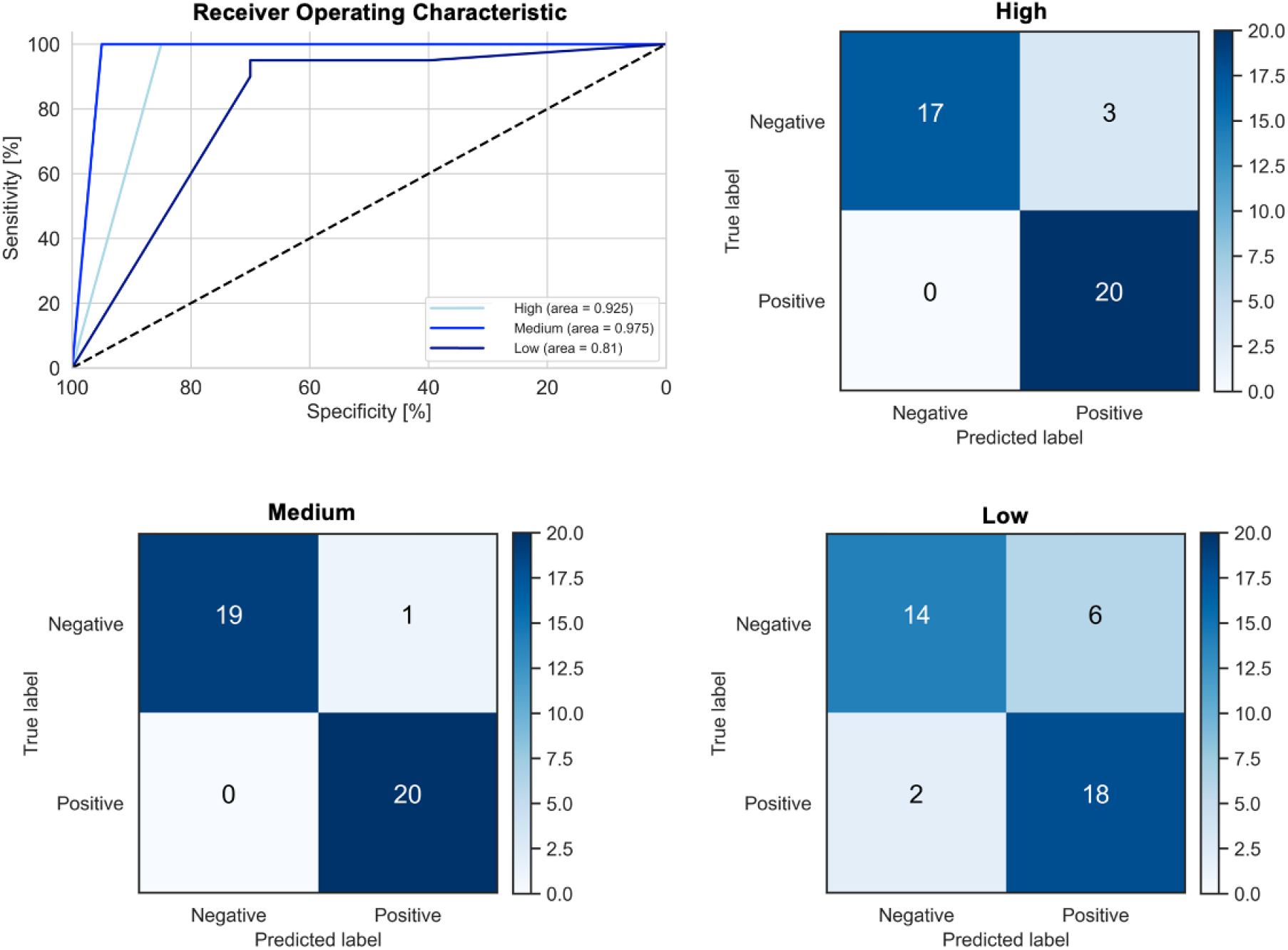
Receiver operating characteristic curves for positive versus negative HN-CT scan discrimination comparing three resampled image resolutions (High – 1.0 mm, Medium – 1.5 mm, and Low – 2.0 mm) and their corresponding confusion matrices.

## Discussion

The incidence of HPV-associated OPC has risen in recent decades and is projected to continue to increase during the next 30 years ^18^. Compared to HPV-negative OPC, HPV-associated OPC has been found to have higher rates of clinical and pathological lymph node involvement ^19^. Additionally, lymph node metastases in HPV-associated OPC are characterized by several distinct features on clinical imaging including cystic composition, and matted conglomeration ^20^. The acquisition of planning HN-CT scans is germane to the radiotherapy treatment workflow. Intravenous iodinated contrast may be administered during the radiotherapy simulation to enhance vascular visibility and soft tissue contrast, thereby facilitating lymph node delineation and manual target volume segmentation ^21,22^.

Patient anatomical and tumor characteristics on medical imaging can be harnessed to automate the process of target volume segmentation for radiotherapy planning. More specifically, DL-CNNs can be used to model complex non-linear relationships in radiation oncology training datasets and make segmentation predictions on unseen HN-CT scans acquired during radiotherapy simulation ^23^. Cardenas et al. used HN-CT scans and their respective, physician-approved contours from 71 patients with head and neck cancers to train, validate, and test a DL-CNN in lymph node clinical target volume (CTV) auto-segmentation. The DL-CNN achieved a DSC of 0.89 for auto-segmented CTVs of neck levels II-V. Additionally, physician review of an independent dataset of 32 HN-CT scans found that over 99% of the DL-CNN auto-segmented lymph node CTVs were either sufficient for clinical use or required minor revisions ^24^.

We designed a DL-CNN using a residual U-Net, a recognized neural network architecture for medical image segmentation ^13,15,23^. Using supervised learning and contrast-enhanced HN-CT scans with corresponding ground-truth masks as inputs, we implemented a patch-based approach to train the DL-CNN to auto-segment involved lymph nodes for patients with HPV-associated OPC. As radiographically occult lymph nodes can be identified on surgical specimens for upward of 50% of patients with head and neck cancers following neck dissection, we confirmed that all radiographically abnormal lymph nodes corresponded to pathologically-involved lymph nodes and that no additional, pathologically-involved were present on surgical histopathology^25,26^.

The role of DL-CNN in the auto-segmentation of head and neck primary tumors on medical imaging has been widely explored ^15,27^. However, studies on auto-segmentation of involved lymph nodes of the head and neck are limited. Bielak et al. investigated the impact of various magnetic resonance imaging sequences on auto-segmentation of lymph nodes and found a maximum DSC of 0.58 ^28^. Similarly, Wang et al. integrated the extraction of various imaging features into a DL-CNN and achieved a mean DSC score of 0.94 for the highest performing model ^29^. As computed tomography scans are acquired during the radiotherapy planning process, we chose to use contrast-enhanced, diagnostic HN-CT scans for the training of our DL-CNN. In order to evaluate the generalization capacity of the DL-CNN auto-segmentation model on unseen data, we split the dataset using 80% for training/validation and 20% for testing. In the validation phase, we found that the DL-CNN achieved median DSC and volume similarity scores of at least 0.90 and 0.95, respectively. When tested on unseen data, the DL-CNN was notable for median consensus segmentation mask scores of 0.92 for DSC and 0.97 for volume similarity. Moreover, the DL-CNN was able to successfully identify node positive HN-CT scans, with an AUC of 0.98. These results suggest that our DL-CNN may be used to perform auto-detection and auto-segmentation of involved lymph nodes as part of the radiation oncology treatment planning workflow with a high degree of fidelity and without the need for additional imaging studies.

There are several limitations to our study. We included patients with HPV-associated OPC who had undergone surgical resection of the primary tumor and lymph node dissection. As this cohort reflects a patient population with early-stage disease, it is possible that our results may not be fully generalizable to patients with more locoregionally advanced disease, including greater than 3 or more radiographically involved lymph nodes and/or radiographic evidence of extranodal extension. Furthermore, our DL-CNN was trained, validated, and tested on contrast-enhanced HN-CT scans. Our findings represent the results of a small cohort of HN-CT scans obtained at a single institution. Therefore, additional studies are needed for external validation of the model in a larger dataset of HN-CT scans performed at other institutions, with and without the presence of intravenous contrast.

## Conclusion

Patients diagnosed with HPV-associated OPC are often found to have clinical evidence of lymph node involvement at the time of diagnosis. Manual segmentation of radiographically involved lymph nodes is an integral part of treatment planning for those patients dispositioned to definitive radiotherapy. Here we have presented a DL-CNN that can be used to automate the process of lymph node detection and segmentation for these patients with a high degree of fidelity. Future studies on the validation of the DL-CNN on larger external datasets of HN-CT scans, on HN-CT scans acquired without contrast, and HN-CT scans pertaining to patients with surgically unresectable disease are necessary to further clarify the role of the DL-CNN in the routine radiation oncology workflow.

## Data Availability

All data produced in the present study are available upon reasonable request to the authors.

