## Supplementary Materials for "Auto-Detection and Segmentation of Involved Lymph Nodes in HPV-Associated Oropharyngeal Cancer Using a Convolutional Deep Learning Neural Network"

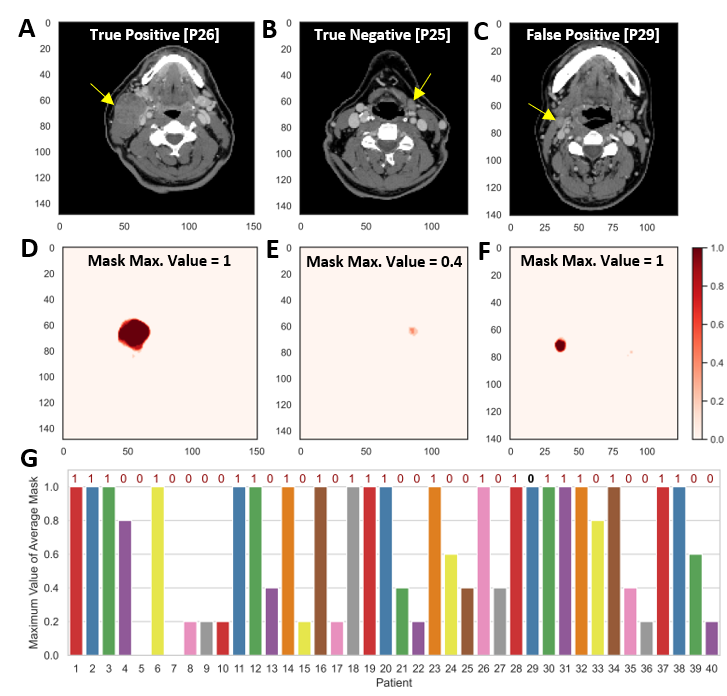

**Figure S1:** Individual lymph node detection results on test set. CT images (A) with a metastatic lymph node and truly identified by the model (True Positive – patient 26), (B) without metastatic lymph nodes and truly identified by the model (True Negative – patients 25), and (C) without metastatic lymph nodes and falsely identified by the model (False Positive – patients 29). Examples of the average masks produced by combining the 5-fold cross validation predicted segmentations on a voxel by voxel basis for (D) True Positive, (E) True Negative, and (F) False Positive cases. (G) A bar plot showing the maximum values of the average mask for the 40 test patients with possible values of (0, 0.2, 0.4, 0.6, 0.8, and 1.0); threshold for prediction was set at 0.8 (≤0.8=0, >0.8=1). The true labels of the patients are given above the bars (1 for positive and 0 for negative).
